# From Blurry to Brilliant: HAGAN, a Hybrid Attention GAN for Remote OCT Image Enhancement

**DOI:** 10.64898/2026.02.23.26346915

**Authors:** Roya Arian, Euan J. Allen, Max Tyler, Raheleh Kafieh

**Affiliations:** Department of Engineering, Durham University, South Road, Durham, DH1 3LE, UK; Siloton Limited, 1 Cathedral Square, Bristol, BS1 5DD, UK

**Keywords:** Optical coherence tomography (OCT), Patient-operated OCT Images, Image enhancement, Generative adversarial networks (GAN), Attention mechanisms, Attention Gates

## Abstract

Regular optical coherence tomography (OCT) is essential for early retinal disease detection, but frequent clinic visits burden patients/healthcare systems. Remote OCT, including home-based imaging, enables more frequent monitoring but introduces variability in image quality due to untrained operation, uncontrolled environments, and cost- and portability-driven hardware constraints. To maximise OCT deployment, hardware and software must overcome these challenges. Here, we focus on the software component by developing a physics-informed simulation framework—Siloton’s simulation software—that models remote OCT acquisition by introducing physically accurate noise and acquisition artifacts into high-quality OCT images, thereby generating low-quality images that mimic sub-optimally acquired patient-operated scans.

Building on this data, we propose **HAGAN**, a Hybrid Attention Generative Adversarial Network designed to enhance low-quality OCT images. The model is developed progressively from a baseline U-Net to an adversarial framework with hybrid-attention. The best-performing U-Net backbone, EfficientNet-B1, identified through systematic evaluation and ablation studies, is adopted as the generator, incorporating attention gates in skip-connections and self-attention in the decoder, and paired with a VGG19-based discriminator. Training uses a multi-objective loss combining pixel, structural, perceptual, edge, and adversarial terms.

Experiments demonstrate that HAGAN consistently outperforms baseline and state-of-the-art methods across standard enhancement metrics and a clinically-relevant retinal layer segmentation downstream-task, improving visual quality while preserving diagnostically meaningful structures. The model also shows strong robustness to progressively increasing noise/artifact levels, maintaining stable performance even under severe degradation. These results highlight the potential of HAGAN to support reliable enhancement for future patient-operated OCT monitoring and effective remote retinal disease management.

**Highlights:**

- Enhancing the quality of sub-optimal OCT images to support remote retinal monitoring and reduce the need for frequent referrals to clinical imaging centers
- Proposing **HAGAN**, a hybrid attention generative adversarial network for enhancing OCT images acquired using the remote OCT devices
- Hybrid attention design combining attention gates and self-attention to preserve fine retinal details while maintaining global anatomical consistency
- Adversarial learning framework improving perceptual realism and preservation of diagnostically relevant retinal structures in low-quality OCT images
- Clinical relevance validated through downstream retinal layer segmentation, confirming preservation of diagnostically important structures
- Demonstrating robustness to progressively increasing noise and artifact levels, where the proposed model maintains stable reconstruction behavior even under severe degradation conditions

## 1. Introduction

Optical coherence tomography (OCT) is a non-invasive imaging modality that enables high-resolution assessment of retinal microstructures and plays a central role in the detection and longitudinal monitoring of major ocular diseases, including diabetic eye disease, age-related macular degeneration, and glaucoma [1, 2, 3, 4]. Given the global burden of vision-threatening retinal diseases, regular OCT monitoring enables early intervention; however, widespread delays in follow-up care continue to place patients with chronic eye disease at risk of preventable sight loss [5, 6, 7].

Despite its clinical importance, conventional OCT imaging remains predominantly hospital-based, requiring specialized equipment and trained operators. This care model places a substantial burden on patients and healthcare systems, as frequent hospital visits are often necessary for disease monitoring—an especially challenging requirement for older individuals. These pressures are further exacerbated by aging populations and the increasing prevalence of chronic eye disease worldwide [8].

In response, global health strategies increasingly prioritise remote and home-based diagnostics; the World Health Organization explicitly promotes digital health, telemedicine, and remote monitoring to decentralise care and improve equity, particularly for older and underserved populations [9, 10].

Recent advances in compact optics and system design have enabled the development of portable, remote OCT devices that aim to facilitate frequent patient-operated retinal imaging outside clinical environments [11, 12, 13] . For example, Siloton has pioneered photonic-chip OCT technology, becoming the first commercial organization to capture retinal images of human subjects using a photonic chip-based OCT device and demonstrating the feasibility of miniaturized imaging outside traditional OCT hardware constraints [14]. While these systems offer substantial practical advantages, factors such as in-dependent user operation, more challenging environmental settings, and potential technical hardware challenges, may change the achieved image quality, particularly with patients that have additional needs (e.g. co-morbidities). Noise, reduced resolution, and structural distortions can, in the worst case, compromise diagnostic accuracy and clinical confidence [15, 16]. Ensuring reliable image quality under sub-optimal imaging conditions therefore remains a critical barrier for equitable-access of remote OCT for the entire patient population.

To reliably study and mitigate image quality degradation in remote OCT acquisition, a simulation framework capable of generating realistic low-quality data from high-quality clinical scans and producing paired datasets suitable for supervised deep learning is required. Simulation-based approaches enable a systematic and cost-effective study phase, allowing controlled analysis and rapid model development prior to real-world experiments, which are substantially more expensive and logistically constrained [17]. To this end, we developed a Siloton simulation framework that converts high-quality hospital OCT scans into simulated lower-quality outputs by modeling the underlying device physics and optical characteristics of a remote OCT system, including clinically relevant degradations such as variations in coherence properties, component scattering, and noise characteristics. Importantly, the simulation is configurable, enabling the generation of a spectrum of image qualities to reflect different hardware limitations and human-factor influences.The resulting paired OCT datasets are then used to train and evaluate the proposed deep learning–based image enhancement model.

Deep learning–based image enhancement methods have demonstrated strong potential for improving degraded OCT images, with encoder–decoder architectures, transfer learning, and adversarial learning frameworks achieving notable gains over traditional signal-processing approaches [18, 19]. However, many existing methods prioritise pixel-wise fidelity and global intensity statistics, often producing over-smoothed outputs that obscure fine retinal structures essential for clinical interpretation [20]. Moreover, prior work often focuses on isolated architectural components without explicitly balancing local detail preservation, global structural consistency, perceptual realism, and downstream clinical relevance [21, 22].

To address these limitations, we propose HAGAN, a Hybrid Attention Generative Adversarial Network for robust enhancement of remote OCT images. The proposed framework integrates complementary local and global attention mechanisms within an adversarial learning paradigm to preserve fine retinal details while maintaining global anatomical consistency and perceptual realism, with anatomical structure preservation assessed through a downstream retinal layer segmentation task [23, 24, 21]. By explicitly balancing pixel-level fidelity with structural and perceptual constraints, HAGAN mitigates over-smoothing effects associated with distortion-driven optimization and enhances the clinical interpretability of reconstructed images [20, 25]. Consequently, HAGAN enables reliable enhancement of degraded, patient-acquired scans and supports the deployment of clinically robust remote OCT systems, contributing to scalable, equitable, and remote retinal monitoring in line with global digital health strategies.

The remainder of this paper is organized as follows. Section 2 reviews related work on OCT image enhancement. Section 3 presents the progressive model development strategy and the proposed HAGAN framework. Section 4 describes the experimental setup and evaluation strategy. Section 5 reports the results and ablation studies, followed by discussion in Section 6 and conclusions in Section 7.

## 2. Related Work

Although patient-operated and home-based OCT systems have gained increasing attention for remote retinal monitoring and demonstrated promising clinical feasibility, prior studies have primarily focused on diagnostic performance, usability, and clinical workflows rather than algorithmic image enhancement [26, 27, 28]. To date, no machine learning–based methods have been specifically developed to enhance the quality of OCT images acquired in patient-operated remote settings, leaving a clear gap in addressing noise, motion artifacts, and other degradations inherent to unsupervised acquisition.

In contrast, deep learning–based image enhancement for clinical OCT has been extensively studied. Convolutional neural networks models have been widely employed to suppress speckle noise, enhance spatial resolution, and improve overall visual quality of retinal OCT images. Among these approaches, U-Net–based architectures, characterized by encoder–decoder structures with skip connections, remain a dominant paradigm due to their strong ability to denoise while preserving fine anatomical details [29, 30, 31, 32, 33]. Furthermore, several studies have integrated pretrained encoder backbones such as ResNet and DenseNet into U-Net variants to improve feature representation and robustness across heterogeneous imaging conditions,demonstrating consistent performance gains over standard convolutional encoders [34, 35].

Beyond purely convolutional models, Generative Adversarial Networks (GANs) have also been explored for OCT image denoising and super-resolution, where adversarial training enables recovery of high-frequency details and perceptually sharper reconstructions compared to pixel-wise loss–only methods [36, 37, 38]. For example, SiameseGAN incorporates a Siamese twin network within a GAN framework to guide the generator toward producing denoised spectral-domain OCT B-scans that are closer to high-SNR ground-truth images in a learned feature space, improving speckle suppression while preserving structural details [39] . Simultaneous Denoising and Super-Resolution (SDSR-OCT) proposes a conditional GAN that jointly performs speckle noise reduction and spatial super-resolution, demonstrating that adversarial learning can address multiple degradations within a unified reconstruction framework [40]. More recently, a pix2pix-style conditional GAN incorporating PatchGAN-based texture loss has been introduced to encourage reconstructed OCT images to match the textural characteristics of gold-standard averaged scans, achieving improved reconstruction quality on clinical retinal datasets [41, 42] . These GAN-based approaches show strong potential for OCT enhancement, although their evaluation has largely been limited to clinical datasets rather than unsupervised remote imaging scenarios.

More recently, attention-augmented GAN architectures have demonstrated notable success in image enhancement tasks, particularly in natural image super-resolution. Models such as Self-Attention GANs (SAGAN) explicitly incorporate attention mechanisms to selectively emphasize informative spatial regions and model long-range dependencies, leading to improved reconstruction fidelity [43].

In the context of OCT imaging, attention mechanisms have primarily been incorporated into convolutional neural networks to emphasize salient anatomical features [44]. However, to the best of our knowledge, the explicit integration of attention mechanisms within GAN frameworks for OCT image enhancement—particularly for remote OCT imaging, where acquisition-related artifacts may be more pronounced—remains relatively underexplored and has not yet been investigated.

To address this gap, we propose an attention-enhanced GAN framework specifically designed for remote OCT image enhancement, aiming to suppress noise and motion artifacts while preserving clinically relevant retinal structures for reliable downstream analysis.

## 3. Methods

Guided by the findings of Section 2, which establish U-Net–based architectures as a strong and widely adopted baseline for OCT image enhancement [29], we adopted a progressive model development strategy. We first implemented a baseline U-Net architecture to establish a reference for reconstruction performance, leveraging encoder–decoder skip connections to preserve structural information and fine-scale anatomical details [45].

Building on prior evidence that advanced convolutional backbones can improve feature representation and robustness in U-Net–based OCT models [34, 35], we integrated several state-of-the-art (SOTA) encoders—including VGG16, VGG19, ResNet-18, DenseNet, ConvNeXt, and EfficientNet-B1—into the U-Net framework to identify the most effective configuration [46, 47]. The best-performing architecture was then adopted as the generator within a generative adversarial network (GAN) framework to exploit adversarial learning and enhance perceptual quality, consistent with the established role of U-Net architectures as effective generators in modern generative models [48].

Finally, we integrated attention mechanisms into the generator’s decoder. Specifically, attention gates were applied at each skip connection to selectively filter encoder features before fusion [49, 23, 30], while self-attention modules were interleaved within the decoder stages to capture long-range dependencies [50]. This hybrid design forms the basis of our proposed model for OCT image enhancement.

### 3.1. Baseline and Progressive Development

We began by implementing a basic autoencoder and introducing skip connections that bridged corresponding encoder and decoder layers, thereby forming a U-Net–style architecture. These skip connections enabled the preservation of both low-level spatial details and high-level semantic features [45]. This design is particularly important for OCT imaging, where subtle textures and fine structural boundaries have to be maintained to achieve clinically meaningful reconstructions [51]. The encoder comprised four convolutional blocks with increasing channel depths (64, 128, 256, and 512), while the decoder mirrored this structure using transposed convolutions.

As the next step in refining the architecture, we systematically evaluated several state-of-the-art convolutional backbones as the encoder of the U-Net to identify the most effective feature representation strategy. To adapt these backbones for use within an encoder–decoder framework, each architecture was decomposed into sequential stages that produce feature maps at progressively deeper levels of abstraction. In the case of VGG16 and VGG19, the convolutional layers were grouped according to their pooling stages, yielding feature maps at five distinct resolutions. For ResNet-18 and DenseNet, the networks were partitioned into their residual or dense blocks, respectively, thereby exposing hierarchical features ranging from low-level edge patterns to high-level semantic representations. For ConvNeXt, the model was divided into its four convolutional stages, consistent with the original hierarchical design. Similarly, EfficientNet-B1 was decomposed into sequential blocks corresponding to progressively deeper feature maps. This decomposition exposed intermediate features at multiple resolutions, which were used to establish skip connections between the encoder and decoder, thereby preserving both low-level structural details and high-level semantic information.

In all backbone encoders, the same training configuration was consistently applied across all models to ensure identical experimental conditions and a fair comparison. Specifically, the first three stages—comprising generic low-level filters pretrained on ImageNet—were frozen, while the deeper blocks were fine-tuned to capture OCT-specific structures. This approach preserves stable low-level representations while enabling the higher layers to adapt to the specialized task of OCT image enhancement[52, 53].

### 3.2. Proposed Model

To overcome the tendency of purely reconstruction-based networks to generate over-smoothed images, we adopted a GAN framework [49, 54]. The winning encoder–decoder architecture from the previous step was employed as the generator, while a VGG19-based discriminator was introduced to enforce perceptual realism by distinguishing between real and generated OCT images. Adversarial training complemented the reconstruction losses, encouraging the generator to produce sharper and more visually convincing results. This setup laid the foundation for the subsequent integration of attention mechanisms. Finally, we enhanced the generator by integrating attention mechanisms into the decoder. Attention gates were applied at each skip connection to selectively filter encoder features before fusion, suppressing irrelevant responses and highlighting task-relevant structures. In parallel, self-attention modules were interleaved within the decoder stages after upsampling to capture long-range spatial dependencies that convolutional layers alone cannot model. This hybrid design, combining local gating with global self-attention, constitutes our proposed model, termed HAGAN, for OCT image enhancement.

In the generator of the HAGAN, self-attention modules are selectively inserted into specific upsampling layers of the decoder to capture long-range dependencies and enrich the contextual representation. The placement of these modules was determined empirically, based on observed improvements in reconstruction quality. The resulting self-attended decoder features—when available, or the raw decoder features otherwise—serve as gating signals in the subsequent attention gates (AGs), which are applied at every skip connection. Each gate integrates decoder guidance with the corresponding encoder feature map to compute spatial attention coefficients, thereby suppressing irrelevant activations while preserving task-relevant structures. Importantly, the decoder pathway itself remains unchanged, while the encoder skip features are modulated by the gate before fusion. Finally, the decoder features and gated encoder features are concatenated, ensuring that only filtered, context-aligned skip information contributes to the reconstruction process.

Consequently, at each decoder stage *i*, the proposed hybrid attention mechanism comprises: (1) self-attention applied to the decoder feature, (2) an attention gate operating on the encoder skip feature, and (3) fusion of the decoder and gated encoder features for hybrid attention. The overall architecture of the proposed HAGAN is illustrated in Figure 1.

**Figure 1:**
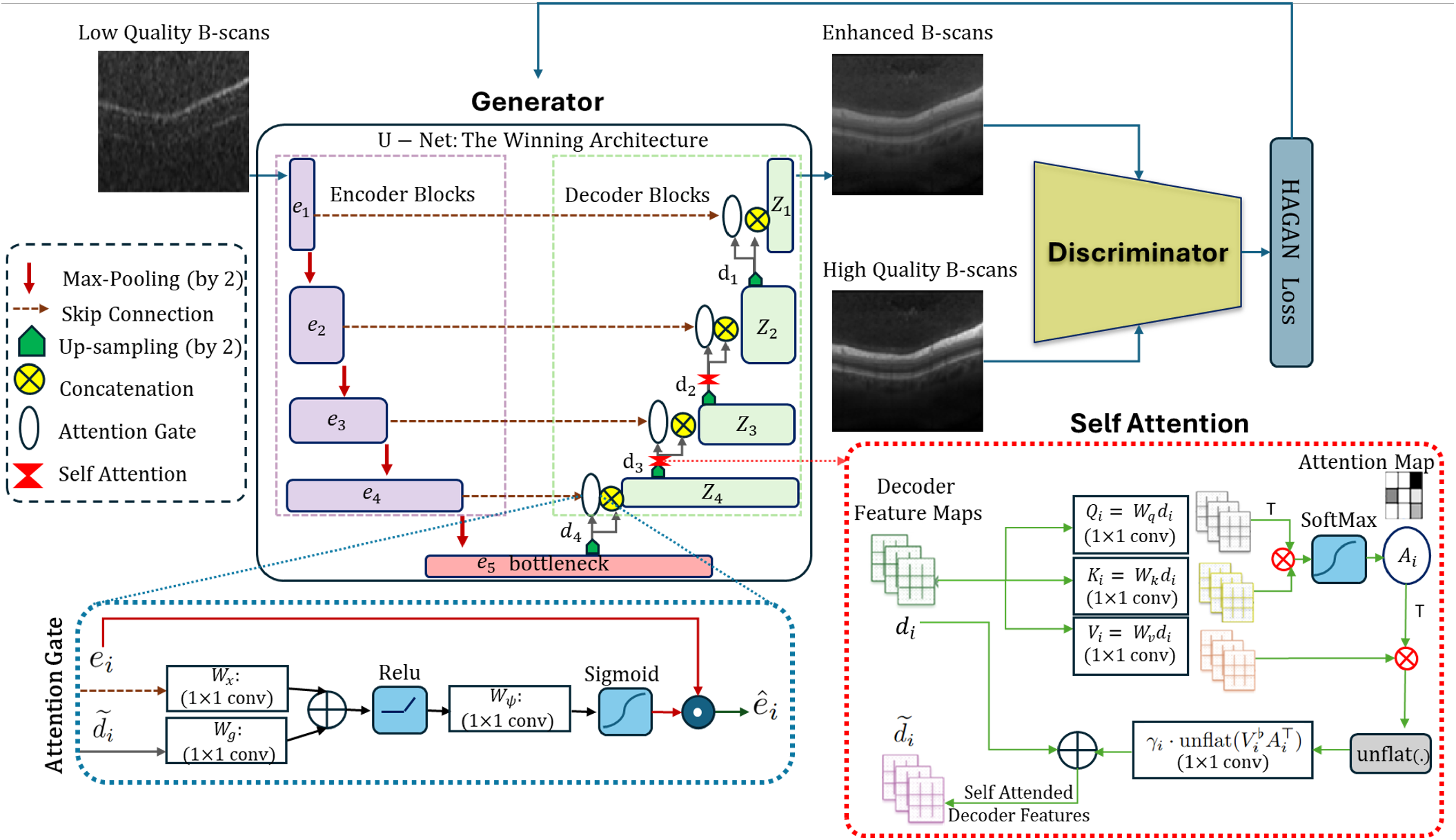
Overview of the proposed HAGAN architecture. The encoder extracts multi-scale features *e*_*i*_, which are filtered by attention gates at each skip connection. Self-attention modules are inserted at selected decoder stages after upsampling to capture long-range dependencies. At each stage, decoder features and gated encoder features are fused to form the hybrid representation, yielding the final enhanced OCT B-scans. **Definitions:** *e*_*i*_: encoder feature at stage *i*; *z*_*i*_: decoder feature at stage *i*; *d*_*i*_: raw decoder feature before self-attention; 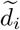: self-attended decoder feature; *ê*_*i*_: gated encoder feature; *A*_*i*_: attention map from self-attention; *γ*_*i*_: learnable scalar weighting the self-attention residual; ⊙: element-wise multiplication; ⊕: element-wise addition; ⊗: matrix multiplication (used in self-attention); T: transpose.

### 3.2.1. Self-Attention on the Decoder Stream

Given the decoder input at stage *i*:

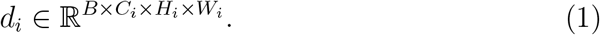

we compute query, key, and value projections:

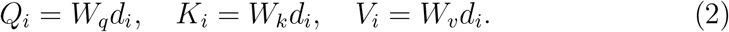

Here, *B* denotes the batch size, and *C*_*i*_ represents the number of channels in the feature map at stage *i*. The spatial dimensions are given by *H*_*i*_ and *W*_*i*_, corresponding to the height and width, respectively. The decoder feature map at stage *i* is denoted by *d*_*i*_. The learnable projection matrices 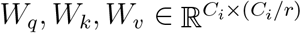 are implemented as 1× 1 convolutions that map the input feature map into query, key, and value embeddings, where *r* is the channel reduction ratio. The resulting tensors 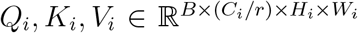 correspond to the query, key, and value representations derived from *d*_*i*_.

Flattening the spatial dimensions (*N* = *H*_*i*_*W*_*i*_), the attention map is:

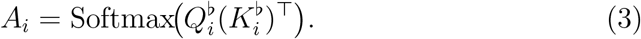

and the self-attended decoder feature is obtained as:

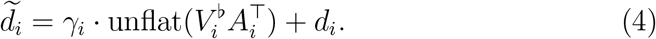

where (·)^*♭*^ denotes flattening to *N* , unflat(·) restores spatial layout, and *γ*_*i*_ is a learnable scalar.

#### 3.2.2. Attention Gate on the Encoder Skip

Let the corresponding encoder feature map be *e*_*i*_. The attention gate computes:

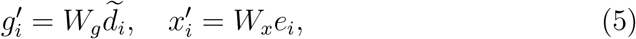

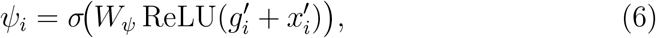

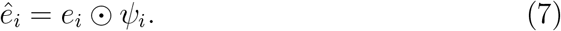

where *W*_*g*_, *W*_*x*_, *W*_*ψ*_ are learnable 1*×*1 convolutions, and *σ*(*·*) is the sigmoid activation. The decoder feature 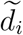serves as the gating signal, while the encoder feature 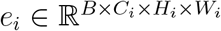 is modulated by the attention coefficients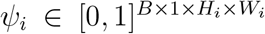. ⊙ denotes element-wise multiplication, where *ψ*_*i*_ is broadcast across the channel dimension of *e*_*i*_ (after spatial sizes are matched if necessary).

#### 3.2.3. Hybrid Attention Fusion

The hybrid decoder output at stage *i* is then:

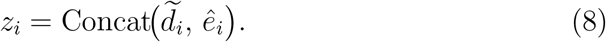

which is passed to the subsequent upsampling block. In compact form:

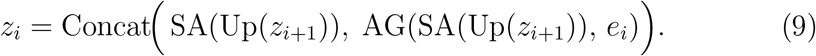

where Up(*·*) denotes transpose convolution upsampling, SA(*·*) is self-attention (applied only at empirically selected layers), AG(*·, ·*) is the attention gate, and Concat(*·*) denotes channel-wise concatenation.

### 3.3. Loss Functions

For all models considered in this study, we employed the same multi-objective loss formulation, combining pixel-wise, structural, perceptual, and edge-preserving terms, with each component addressing a different aspect of the image enhancement task. For HAGAN, we additionally introduced an adversarial component, enabling the generator to benefit from the discriminator’s feedback. The final generator objective is a weighted sum of these components, as detailed below. The definitions of all symbols used in the loss function formulations are provided in Table 1.

**Table 1:** List of Symbols Used in the Loss Functions.

| Symbol | Definition |
| --- | --- |
| $x$ | Low-quality OCT B-scan (input) |
| $y$ | High-quality OCT B-scan (ground truth) |
| $G_\theta(\cdot)$ | Generator network with parameters $\theta$ |
| $\mathbb{E}_{x,y}[\cdot]$ | Expectation over training dataset pairs $(x, y)$ |
| $\ \cdot\ _1, \ \cdot\ _2^2$ | $\ell_1$ and squared $\ell_2$ norms |
| $\mu_p, \mu_q$ | Local means of patches $p$ and $q$ |
| $\sigma_p^2, \sigma_q^2$ | Local variances of patches $p$ and $q$ |
| $\sigma_{pq}$ | Cross-covariance of patches $p$ and $q$ |
| $C_1, C_2$ | Small constants to stabilize the division |
| $\nabla$ | Image gradient operator (for edge loss) |
| $\mathcal{S}$ | Set of selected feature layers |
| $\varphi_\ell(\cdot)$ | Feature map at layer $\ell$ of pretrained VGG19 |
| $w_\ell$ | Weight assigned to perceptual layer $\ell$ |
| $D_\phi(\cdot)$ | Discriminator network with parameters $\phi$ |
| $\lambda_{(\cdot)}$ | Hyperparameters weighting each loss component |

### 3.3.1. Pixel-wise Reconstruction Losses

To enforce fidelity at the pixel level, we employ both *𝓁*_1_ and *𝓁*_2_ penalties:

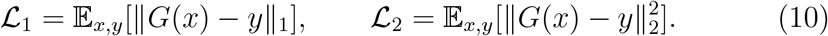

#### 3.3.2. Structural Similarity Loss

The structural similarity (SSIM) term improves perceptual quality by aligning local structures between the generated image *G*(*x*) and the target image *y*:

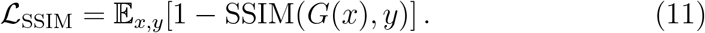

Where the SSIM index between two image patches *p* and *q* is defined as

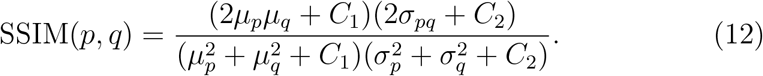

This value lies in [*−*1, 1], with 1 indicating perfect structural similarity.

#### 3.3.3. Edge-preserving Loss

To encourage sharper retinal boundaries, we compute gradient differences between the prediction and ground truth:

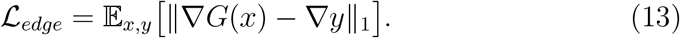

#### 3.3.4. Perceptual Loss

To account for perceptual similarity as perceived by the human visual system, we employ a feature-based loss that measures distances in the representational space of a pretrained network *φ* (e.g., VGG19). Unlike pixel-wise measures that enforce strict intensity correspondence, this approach captures higher-level structural and textural attributes, which are more consistent with human judgments of image quality. Formally, the perceptual loss is defined as:

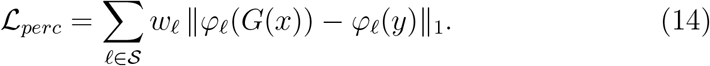

#### 3.3.5. Adversarial Loss

The adversarial term encourages the generator *G* to produce outputs *ŷ* = *G*(*x*) that are indistinguishable from real high-quality OCT images *y*. The discriminator *D* is trained to distinguish between real and generated samples:

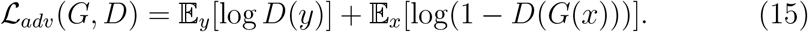

The generator’s adversarial loss is then

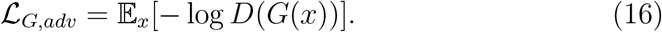

#### 3.3.6. Final Generator Loss

The complete generator objective is a weighted combination:

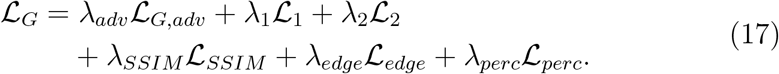

The discriminator is trained by minimizing *ℒ*_*adv*_(*G, D*).

For all other models considered in this study, the same multi-objective formulation was employed, excluding the adversarial term (i.e., *λ*_adv_*ℒ*_*G*,adv_).

## 4. EXPERIMENTS

### 4.1. Dataset

In this study, a total of 5000 B-scans from 100 patients were used for training and validation with a 90:10 split, and an additional independent set of 1000 B-scans from 20 patients served as the unseen test dataset. All B-scans were macular spectral-domain OCT (SD-OCT) images acquired using the Heidelberg Spectralis HRA+OCT system (Heidelberg Engineering, Heidelberg, Germany) at Noor Ophthalmology Hospital, Tehran, Iran. The dataset consisted exclusively of healthy Iranian subjects [55].

Eligible participants were between 18 and 89 years of age. Written informed consent was obtained from all participants, and the study was conducted in accordance with institutional ethical standards and the Declaration of Helsinki. The acquired OCT volumes varied in size from 512× 496× 19 to 512 ×496 ×120 voxels along the *x, y*, and *z* axes, respectively.

All B-scans were then resized to 256 ×256× 3 to preserve structural detail while remaining compatible with ImageNet-pretrained backbones, which can accept arbitrary input sizes. The single-channel OCT images were replicated across three channels, and all pixel intensities were normalized to the range [0, 1] by dividing by 255.

### 4.2. Simulation of Remote OCT Data

Due to the limited availability of real remote OCT data and the absence of corresponding high-quality reference images acquired using the same device as the low-quality scans, we generated a synthetic paired dataset through simulation. Specifically, we developed a proprietary Siloton simulation software that leverages in-hand high-quality clinical OCT images to emulate the image formation process of a remote OCT device under arbitrary performance conditions. The software incorporates key acquisition parameters that model the optical and electronic characteristics of the system, transforming high-quality images into variable low-quality counterparts. The degradation process is tunable, enabling the generation of a wide range of output qualities that reflect variability across different OCT devices, acquisition conditions, and patient-specific factors. The resulting simulated images exhibit noise characteristics, resolution degradation, and acquisition artifacts consistent with those observed in real outputs. Through this controlled degradation process, each simulated low-quality image remains directly paired with its original high-quality source, yielding a supervised dataset suitable for training and evaluating OCT image quality enhancement models under reproducible and well-defined conditions.

Figure. 2 presents representative high-quality OCT images alongside their paired simulated low-quality versions, produced with the Siloton simulation software to mimic the characteristics of a poorly operating OCT device.

**Figure 2:**
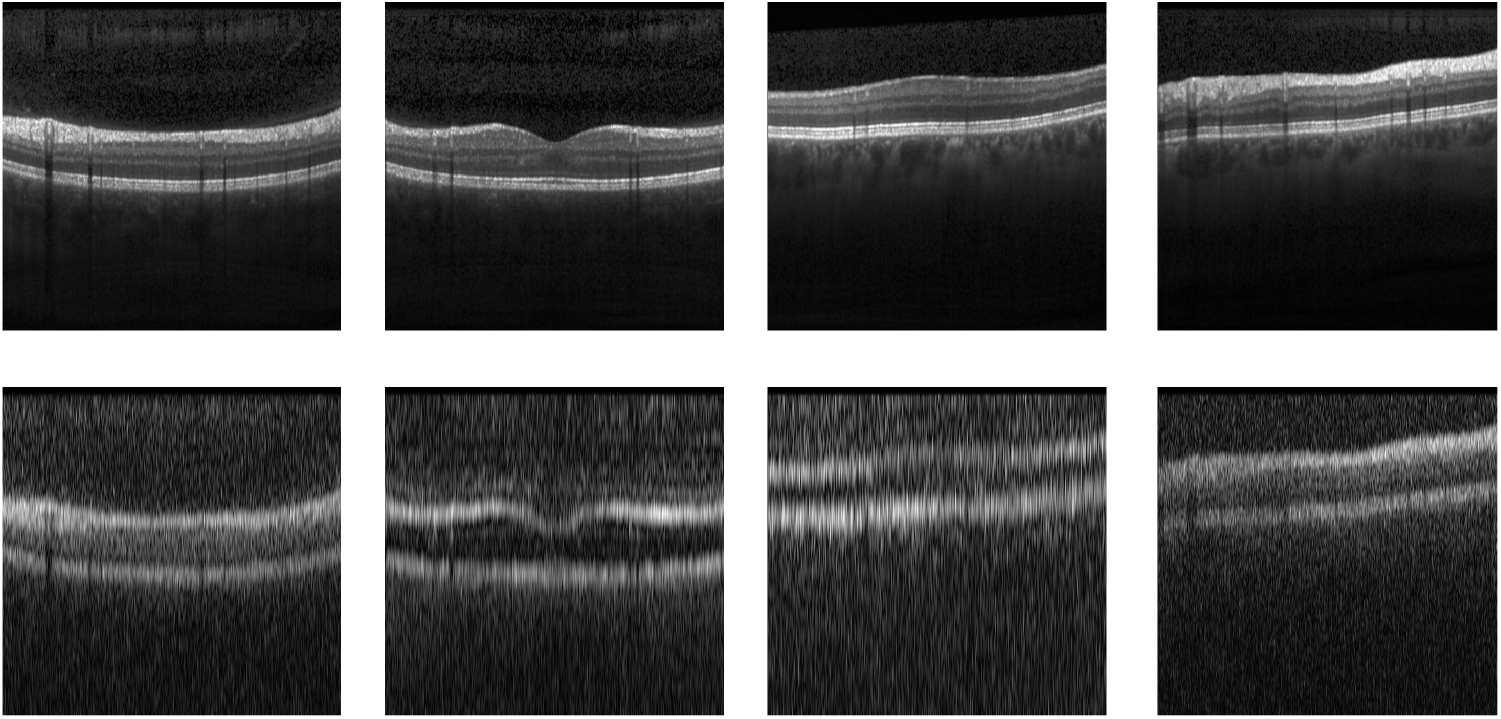
Representative examples of high-quality OCT B-scans (top row) and their paired simulated low-quality counterparts (bottom row), generated using the Siloton simulation software. The simulated images reproduce the noise patterns, resolution limitations, and acquisition artifacts characteristic of a remote OCT device with significantly comprimised imaging performance

### 4.3. Implementation details

All experiments were conducted on a workstation with 128 GB of RAM and NVIDIA GeForce RTX 4090 GPUs (24 GB) running CUDA 12.6. The implementation was developed in Python 3.10.16 on Linux, using PyTorch and a standardized research template to ensure reproducibility.

To balance the contribution of different loss terms, we empirically determined the weighting coefficients for each loss function and fixed them across all models. The only exception was the HAGAN model, in which we additionally incorporated an adversarial loss term with a dedicated weight. Table 2 summarizes the final values of the loss weights used in our experiments.

**Table 2:** Loss function weights used in our experiments. All models employed the same configuration, except HAGAN which additionally included the adversarial loss term.

| Loss Term | Weight ($\lambda$) |
| --- | --- |
| $\lambda_{l1}$ | 0.5 |
| $\lambda_{l2}$ | 1.7 |
| $\lambda_{ssim}$ | 0.2 |
| $\lambda_{edge}$ | 0.5 |
| $\lambda_{perc}$ | 0.002 |
| $\lambda_{adv}$ (HAGAN only) | 0.005 |

We subsequently employed the Optuna framework to optimize the hyperparameters of the HAGAN model [56, 57]. Optuna’s Bayesian optimization with pruning was used to efficiently explore the search space, tuning parameters such as the learning rate and optimizer configuration for both the generator and discriminator, as well as the number of fully connected layers and the number of neurons per layer in the discriminator. As both the generator and discriminator are based on pretrained CNN architectures, the convolutional layers inherited from these backbones were kept fixed during the optimization process, while only training-related parameters and the fully connected layers of the discriminator were optimized.

The best-performing configuration, determined based on validation performance, was adopted for the final experiments. These tuned hyperparameters are presented in Table 3.

**Table 3:** Tuned hyperparameters of the HAGAN model obtained using Optuna.

| Hyperparameter | Selected Value |
| --- | --- |
| Number of discriminator layers | 1 |
| Number of neurons per layer | 78 |
| Generator optimizer | AdamW |
| Generator learning rate | $8.43 \times 10^{-4}$ |
| Discriminator optimizer | Adam |
| Discriminator learning rate | $1.20 \times 10^{-5}$ |

### 4.4. Evaluation Metrics

To evaluate the performance of the different frameworks, we employed several widely used metrics that capture both pixel-level fidelity and perceptual quality.

### 4.4.1. Standard Image Enhancement Metrics

The following standard metrics were used in our experiments:

- Mean Absolute Error (MAE): Measures the average absolute difference between predicted and reference images; lower values indicate more accurate reconstructions.
- Mean Squared Error (MSE): Measures the average squared difference between predicted and reference images, emphasizing larger errors.
- Peak Signal-to-Noise Ratio (PSNR): Measures reconstruction fidelity by comparing the signal strength to the level of distortion; higher values indicate better quality.
- Structural Similarity Index (SSIM): Measures luminance, contrast, and structural similarity between two images; values closer to 1 reflect higher perceptual quality.
- Learned Perceptual Image Patch Similarity (LPIPS): Measures perceptual similarity based on deep feature representations; lower values correspond to images that are more visually similar to the reference.

Together, these metrics provide a comprehensive evaluation of both fidelity and perceptual realism across the tested models.

#### 4.4.2. Downstream Evaluation via Segmentation

We did not rely solely on these quantitative metrics, as there is currently no universally accepted standard for evaluating image enhancement [20]. Different models can perform better on some metrics while worse on others, and there are no well-defined threshold values that directly indicate clinical usefulness. To provide a more meaningful evaluation, we therefore considered a downstream task of clinical relevance: segmentation of retinal layers within OCT B-scans [58].

Layer delineation is particularly important for ophthalmologists, as the distinction between retinal layers underpins the diagnosis and monitoring of a wide range of ocular diseases [57, 59]. To this end, we employed a pretrained segmentation model capable of delineating nine distinct retinal layers and assessed whether the reconstructed images preserved sufficient structural information to accurately recover these boundaries [60]. This employed segmentation model follows an Attention U-Net architecture with a pretrained CNN encoder and a decoder with skip connections enhanced by attention gates, enabling the network to focus on relevant retinal structures [61]. The model was trained on high-quality OCT B-scans with expert annotations of nine retinal layers from the HCIRAN dataset [55], acquired using a Heidelberg Spectralis device. Importantly, the segmentation network was trained only on these high-quality scans and not on the low-quality or reconstructed images used in this study. In the original framework, segmentation performance was evaluated using the Dice Similarity Coefficient (DSC) and the Mean Absolute Distance (MAD) to measure boundary localization accuracy [62].

We applied the segmentation model to the low-quality inputs, the reconstructed images generated by different enhancement frameworks, and the ground-truth high-quality B-scans in order to assess the impact of image enhancement on segmentation performance. To enable a layer-wise comparison, mean squared error (MSE) was computed between the predicted probability maps and the corresponding ground-truth boundaries. This boundary-focused metric evaluates how accurately the predicted layer probabilities align with the ground-truth retinal layer boundaries. The performance of the segmentation model on a representative high-quality image is illustrated in Figure 3.

**Figure 3:**
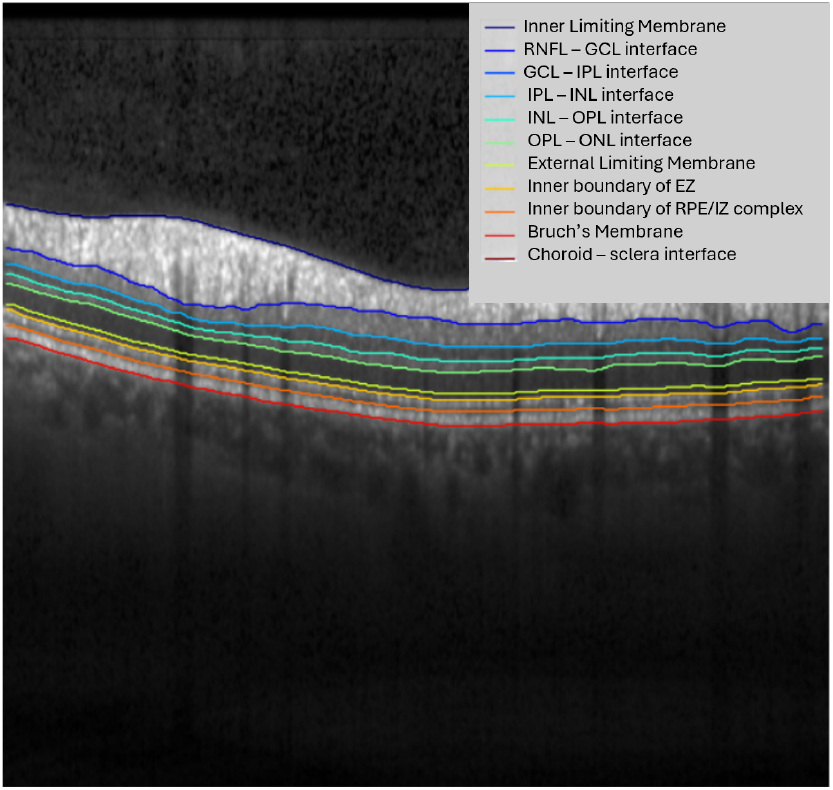
Performance of the pretrained segmentation model on a representative high-quality OCT B-scan. The model is capable of delineating nine distinct retinal layers, which are of critical importance for ophthalmological diagnosis.

## 5. RESULTS

### 5.1. U-Net Architectures Comparisons

We conducted a systematic comparison of U-Net–based architectures reported in the literature [29], focusing on the impact of different encoder backbones on OCT image enhancement performance. Specifically, we evaluated U-Net variants incorporating VGG16 and VGG19 [63], ResNet18 [34], DenseNet121 [64], ConvNeXt [65], and EfficientNet-B1 [66]. Each backbone was integrated within a consistent U-Net framework to enable a fair assessment of its ability to extract hierarchical and multi-scale features relevant to OCT enhancement.

Quantitative results summarized in Table 4 demonstrate that raw low-quality inputs suffer from markedly degraded performance, highlighting the severe impact of acquisition-related artifacts on both perceptual image quality and anatomical fidelity. In contrast, reconstruction-based enhancement effectively restores structural information, enabling reliable downstream analysis. Across standard image enhancement metrics, EfficientNet-B1 consistently outperformed other encoder backbones, achieving the lowest MAE and MSE alongside the highest PSNR and SSIM, while also producing the most perceptually faithful reconstructions as indicated by LPIPS. Segmentation-based evaluation using layer-wise MSE further confirmed its superiority, with EfficientNet-B1 yielding the most accurate preservation of retinal boundaries.

**Table 4:** Quantitative comparison of different U-Net architectures. Each row is a metric, and columns show model performance. Lower is better for ↓, higher is better for↑ .

| Metric | U-Net[29] | VGG16[63] | VGG19[63] | ResNet18[34] | DenseNet121[64] | ConvNeXt[65] | EfficientNet-B1[66] | LowQ |
| --- | --- | --- | --- | --- | --- | --- | --- | --- |
| **Enhancement Metrics** |  |  |  |  |  |  |  |  |
| MAE↓ | 0.0280 | 0.0258 | 0.0299 | 0.0242 | 0.0264 | 0.0247 | **0.0209** | 0.0587 |
| MSE↓ | 0.0020 | 0.0015 | 0.0019 | 0.0014 | 0.0016 | 0.0014 | **0.0010** | 0.0077 |
| PSNR↑ | 27.22 | 28.37 | 27.49 | 28.83 | 28.35 | 28.53 | **30.16** | 21.55 |
| SSIM↑ | 0.7919 | 0.8079 | 0.7971 | 0.8063 | 0.7889 | 0.8059 | **0.8158** | 0.4406 |
| LPIPS↓ | 0.1649 | 0.1713 | 0.1776 | 0.1936 | 0.2230 | 0.1653 | **0.1584** | 0.5194 |
| **Segmentation Performance (Boundary MSE ↓)** |  |  |  |  |  |  |  |  |
| L0 | 0.001936 | **0.001375** | 0.001793 | 0.001453 | 0.001690 | 0.001544 | 0.001464 | 0.012093 |
| L1 | 0.004830 | 0.003648 | 0.003922 | 0.003670 | 0.004385 | 0.003731 | **0.003242** | 0.014957 |
| L2 | 0.006406 | 0.004618 | 0.004476 | 0.004463 | 0.005175 | 0.004507 | **0.004095** | 0.016610 |
| L3 | 0.007533 | 0.004883 | 0.004629 | 0.004754 | 0.005050 | 0.004749 | **0.004542** | 0.013159 |
| L4 | 0.008217 | 0.004909 | 0.004862 | 0.004939 | 0.005313 | 0.004977 | **0.004462** | 0.013345 |
| L5 | 0.010389 | 0.003857 | 0.003949 | 0.004189 | 0.004874 | 0.005642 | **0.003530** | 0.021043 |
| L6 | 0.011451 | 0.003015 | 0.003308 | 0.003551 | 0.004469 | 0.006517 | **0.002695** | 0.010154 |
| L7 | 0.012916 | 0.003192 | 0.003585 | 0.003954 | 0.004610 | 0.006955 | **0.002699** | 0.013664 |
| L8 | 0.013505 | 0.003507 | 0.004034 | 0.004274 | 0.004808 | 0.007554 | **0.003168** | 0.017587 |
| L9 | 0.007987 | 0.002097 | 0.002464 | 0.002339 | 0.002779 | 0.004393 | **0.001987** | 0.044749 |
| Avg | 0.008517 | 0.003510 | 0.003679 | 0.004159 | 0.004315 | 0.005057 | **0.003189** | 0.017736 |

### 5.2. Ablation Studies on the Optimal U-Net

After selecting EfficientNet-B1 as the best-performing backbone, we conducted a series of ablation studies focusing on the number of frozen encoder stages and the role of skip connections within the EfficientNet–U-Net architecture. First, we evaluated the contribution of skip connections by selectively removing them to quantify their effect on preserving spatial detail and reconstruction quality. Second, as mentioned previously, in the initial experiments with all backbone encoders, including EfficientNet-B1, the first three encoder stages were frozen while only the deeper encoder blocks were finetuned to capture OCT-specific structures. Building on this configuration, we further investigated the optimal degree of fine-tuning by systematically unfreezing an increasing number of encoder stages, starting from a fully frozen encoder (i.e., transfer learning) up to unfreezing four encoder stages. Importantly, the unfrozen stages correspond to the deepest encoder blocks located closest to the latent representation, which are responsible for learning highlevel, task-specific features. The results of these ablation experiments are summarized in Table 5.

**Table 5:** Ablation study on encoder stage freezing and skip connections using EfficientNet-B1 as the backbone. Each row reports a performance metric, while the columns correspond to different ablation configurations. Lower values indicate better performance for metrics marked with *↓*, whereas higher values indicate better performance for metrics marked with*↑*.

| Metric | No Skip | Transfer | 1 Unfrozen | 2 Unfrozen | 3 Unfrozen | 4 Unfrozen | LowQ |
| --- | --- | --- | --- | --- | --- | --- | --- |
| **Enhancement Metrics** |  |  |  |  |  |  |  |
| MAE↓ | 0.0218 | 0.0256 | 0.0206 | 0.0209 | **0.0187** | 0.0191 | 0.0587 |
| MSE↓ | 0.0013 | 0.0016 | 0.0011 | 0.0010 | **0.0009** | **0.0009** | 0.0077 |
| PSNR↑ | 29.07 | 28.13 | 30.00 | 30.16 | **30.86** | 30.69 | 21.55 |
| SSIM↑ | 0.8266 | 0.8210 | 0.8448 | 0.8158 | **0.8529** | 0.8511 | 0.4406 |
| LPIPS↓ | 0.2614 | 0.1671 | **0.1560** | 0.1584 | 0.1608 | 0.1597 | 0.5194 |
| **Segmentation Performance (Boundary MSE ↓)** |  |  |  |  |  |  |  |
| L0 | 0.001939 | 0.002030 | 0.001434 | 0.001464 | **0.001340** | 0.001369 | 0.012093 |
| L1 | 0.004471 | 0.004691 | 0.003400 | 0.003242 | **0.003097** | 0.003136 | 0.014957 |
| L2 | 0.005473 | 0.005792 | 0.004519 | 0.004095 | **0.003918** | 0.004027 | 0.016610 |
| L3 | 0.005730 | 0.006059 | 0.005026 | 0.004542 | **0.004317** | 0.004472 | 0.013159 |
| L4 | 0.005779 | 0.006065 | 0.004900 | 0.004462 | **0.004287** | 0.004458 | 0.013345 |
| L5 | 0.004734 | 0.005812 | 0.004101 | 0.003530 | **0.003371** | 0.003530 | 0.021043 |
| L6 | 0.003657 | 0.005111 | 0.003516 | 0.002695 | **0.002644** | 0.002687 | 0.010154 |
| L7 | 0.003804 | 0.005479 | 0.003593 | **0.002699** | 0.002787 | 0.002743 | 0.013664 |
| L8 | 0.004162 | 0.006238 | 0.004131 | **0.003168** | 0.003272 | 0.003204 | 0.017587 |
| L9 | 0.002329 | 0.004147 | 0.002534 | **0.001987** | 0.001992 | 0.001989 | 0.044749 |
| Avg | 0.004607 | 0.005342 | 0.003863 | 0.003189 | **0.003103** | 0.003284 | 0.017736 |

As shown in Table 5, skip connections consistently improve performance, and unfreezing three encoder stages yields the best overall results compared to configurations with no unfrozen stages or with one, two, or four unfrozen stages. Although unfreezing four encoder stages leads to marginal improvements on certain individual metrics, the configuration with three unfrozen stages achieves superior overall performance while incurring substantially lower computational cost. Accordingly, the EfficientNet–U-Net architecture with three unfrozen encoder stages—specifically the deepest layers near the latent space—and skip connections was selected as the generator for the proposed HAGAN model.

### 5.3. Proposed Hybrid Attention GAN (HAGAN)

As described earlier, the proposed HAGAN employs an EfficientNet-U-Net with three unfrozen encoder stages and skip connections as the generator, while the discriminator is based on a VGG19 backbone. To further enhance performance, hybrid attention mechanisms were incorporated, combining attention gates for local feature refinement and self-attention for capturing global context. The comparative results between the proposed HAGAN and the baseline EfficientNet-U-Net are presented in Table 6, demonstrating the effectiveness of the hybrid attention GAN design.

**Table 6:** Attention ablation within EfficientNet-U-Net (three unfrozen encoder stages) and comparison to the full HAGAN. Each row reports a performance metric, while the columns correspond to different ablation configurations. Lower is better for↓, higher is better for ↑. HAGAN: proposed Hybrid GAN model; SAGAN: self-attention GAN, AGGAN: attention gates GAN

| Metric | GAN (No Attn)[37] | AGGAN | SAGAN [43] | HAGAN (AG+SA) | EfficientNet-U-Net[66] |
| --- | --- | --- | --- | --- | --- |
| **Enhancement Metrics** |  |  |  |  |  |
| MAE↓ | 0.0190 | 0.0188 | 0.0189 | **0.0181** | 0.0187 |
| MSE↓ | 0.0010 | 0.0009 | 0.0009 | 0.0009 | 0.0009 |
| PSNR↑ | 30.46 | 30.55 | 30.50 | 30.83 | **30.86** |
| SSIM↑ | 0.8420 | 0.8437 | 0.8421 | **0.8537** | 0.8529 |
| LPIPS↓ | 0.1612 | 0.1605 | 0.1642 | **0.1542** | 0.1608 |
| **Segmentation Performance (Boundary MSE ↓)** |  |  |  |  |  |
| L0 | 0.001316 | 0.001317 | 0.001328 | **0.001305** | 0.001340 |
| L1 | 0.003327 | 0.002971 | 0.003252 | **0.002969** | 0.003097 |
| L2 | 0.004126 | 0.003849 | 0.004058 | **0.003794** | 0.003918 |
| L3 | 0.004303 | 0.004277 | 0.004226 | **0.004199** | 0.004317 |
| L4 | 0.004225 | 0.004198 | 0.004233 | **0.004164** | 0.004287 |
| L5 | 0.003390 | 0.003337 | 0.003445 | **0.003289** | 0.003371 |
| L6 | 0.002587 | 0.002568 | 0.002663 | **0.002502** | 0.002644 |
| L7 | 0.002651 | 0.002547 | 0.002590 | **0.002474** | 0.002787 |
| L8 | 0.003118 | 0.002933 | 0.003109 | **0.002709** | 0.003272 |
| L9 | 0.001854 | 0.001834 | 0.001914 | **0.001643** | 0.001992 |
| Avg↓ | 0.030897 | 0.002983 | 0.030818 | **0.002905** | 0.003103 |

### 5.4. Ablation Studies on HAGAN

#### 5.4.1. Attention Mechanisms

To isolate the contribution of attention mechanisms, we performed a systematic ablation study on the generator architecture (EfficientNet–U-Net with three unfrozen encoder stages), focusing on attention gates (AG) on skip connections and self-attention (SA) modules. We evaluated four GAN configurations that differ only in the generator’s attention design: (i) no attention, serving as a GAN baseline representative of conventional adversarial OCT enhancement methods [37]; (ii) AG-only (AGGAN), incorporating attention gates on skip connections; (iii) SA-only (SAGAN), employing self-attention modules in the generator [43]; and (iv) a hybrid AG+SA configuration.

As summarized in Table 6, both AGGAN and SAGAN outperform the no-attention baseline. Attention gates primarily improve local structural detail by selectively filtering encoder features, whereas self-attention enhances global coherence by modeling long-range spatial dependencies, consistent with prior SAGAN-based findings [43]. The hybrid AG+SA design achieves the best overall trade-off across perceptual quality (LPIPS), pixel-level fidelity (MAE, MSE, PSNR, SSIM), and segmentation-based evaluation (average layer-wise MSE). Incorporating adversarial training within the proposed HAGAN framework further amplifies these gains, highlighting the complementary benefits of local and global attention mechanisms for structure-preserving OCT image enhancement.

#### 5.4.2. Degradation Levels

To investigate the robustness of the proposed HAGAN framework under progressively degraded imaging conditions, we evaluated the model across multiple levels of simulated noise and artifact corruption. The degradation process was applied iteratively in five stages. At each stage, the previously degraded image was used as the input to the noise-generation model, producing progressively more challenging imaging conditions with increasing noise and artifact severity. The model was progressively fine-tuned on images from each degradation level, allowing it to adapt to increasingly corrupted inputs.

Figure 4 summarizes the results of this experiment. In Fig. 4(a), representative qualitative examples across degradation levels are shown, including the degraded inputs and their corresponding reconstructions produced by HAGAN. As the degradation severity increases, the input images become progressively dominated by noise and artifacts. Nevertheless, the reconstructed outputs consistently recover the underlying retinal structures and preserve key anatomical boundaries.

**Figure 4:**
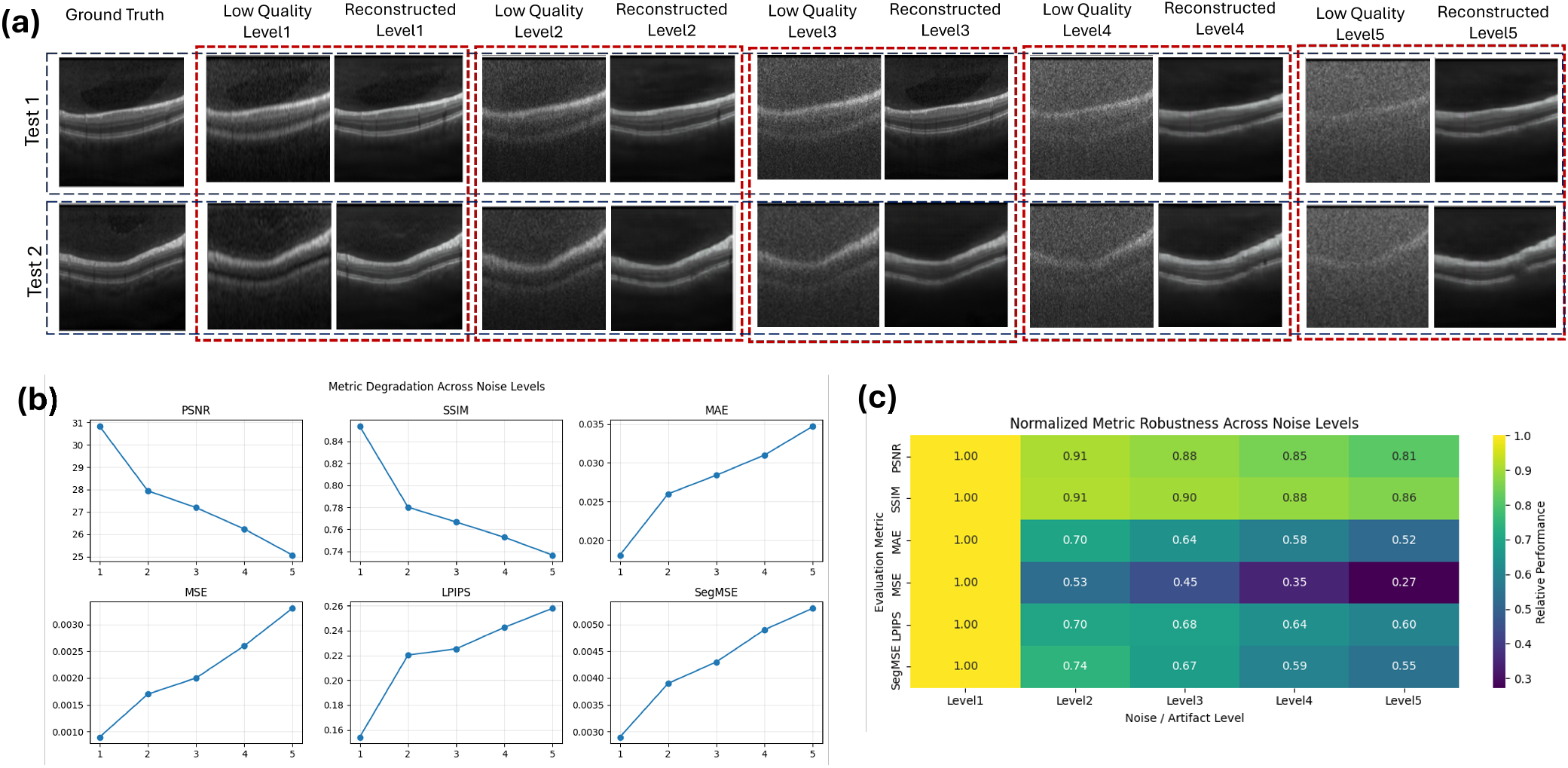
Robustness evaluation of HAGAN across increasing degradation levels. (a): Representative qualitative examples showing progressively degraded inputs and the corresponding reconstructions produced by HAGAN. (b): Quantitative trends of evaluation metrics (PSNR, SSIM, MAE, MSE, LPIPS, and SegMSE) as the degradation level increases. (c): Heatmap of normalized metrics illustrating the relative robustness of the model across noise levels.

The quantitative behavior is illustrated in Fig. 4(b), where the metric plots show the evolution of PSNR, SSIM, MAE, MSE, LPIPS, and segmentation boundary error (SegMSE) as the degradation level increases. As degradation severity increases, the reconstruction task becomes progressively more challenging; nevertheless, HAGAN remains effective in recovering meaningful anatomical structures, with only a gradual reduction in performance under severe corruption.

For easier cross-metric comparison, Fig. 4(c) presents a normalized heatmap of the evaluation metrics across degradation levels. The heatmap highlights that perceptual and structural metrics degrade more gradually than pixel-wise error measures, suggesting that HAGAN preserves structural information even when pixel-level noise becomes significant.

Overall, these results demonstrate that the proposed HAGAN model remains capable of recovering meaningful retinal structures across a wide range of degradation levels, confirming its robustness to severe noise and artifact conditions.

### 5.5. Qualitative Results

In addition to the quantitative improvements, Figure 5 presents representative qualitative examples that illustrate the visual and structural benefits of the proposed approach. As shown, the enhanced images produced by the proposed model exhibit clearer retinal structures and improved continuity of retinal layer boundaries.

**Figure 5:**
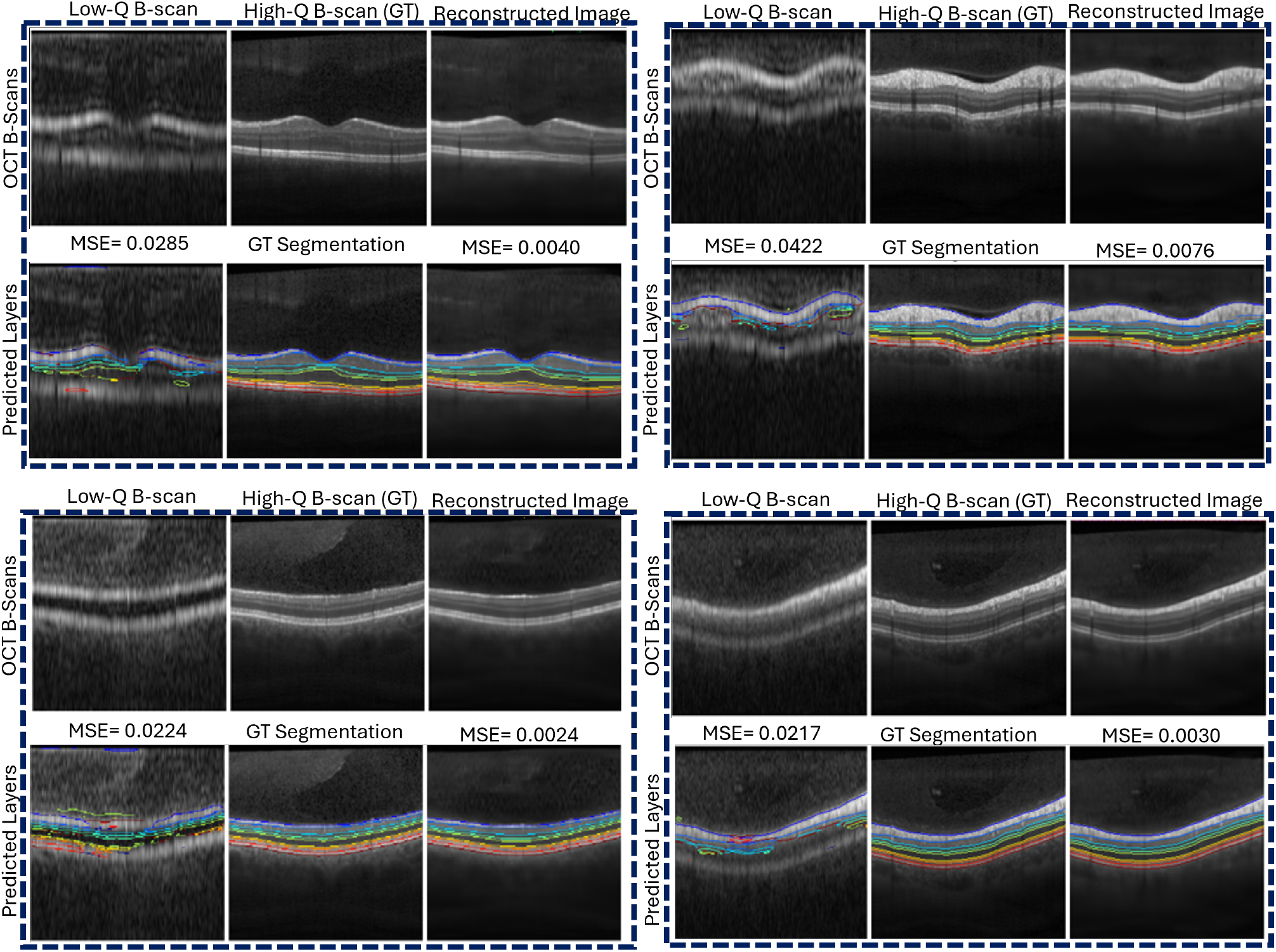
Qualitative results of the proposed Hybrid Attention GAN (HAGAN). For each example, the top row presents, from left to right, the input low-quality OCT B-scan, the corresponding ground-truth high-quality image, and the enhanced output generated by HAGAN. The bottom row displays the associated retinal layer segmentations. Overall, the proposed HAGAN demonstrates improved structural clarity and more consistent delineation of retinal layer boundaries across a range of image qualities.

Furthermore, Figure 6 enables a direct qualitative comparison of the progressive model development employed in this study, from a simple baseline U-Net to the final Hybrid Attention GAN (HAGAN). Specifically, four models are evaluated: (i) a baseline U-Net, (ii) an EfficientNet-U-Net selected as the best-performing U-Net and autoencoder-based architecture, (iii) a GAN model using the winning EfficientNet-U-Net as the generator, and (iv) the proposed HAGAN. The segmentation results demonstrate that the baseline U-Net suffers from local discontinuities and fragmented retinal layer boundaries, particularly in challenging regions. These segmentation inconsistencies are progressively reduced as the model architecture is refined, with improved boundary coherence observed in the EfficientNet-U-Net and further enhanced in the GAN-based formulation. The final HAGAN model yields the most consistent and anatomically coherent retinal layer segmentation, highlighting the benefit of the proposed progressive design strategy and the Final proposed HAGAN.

**Figure 6:**
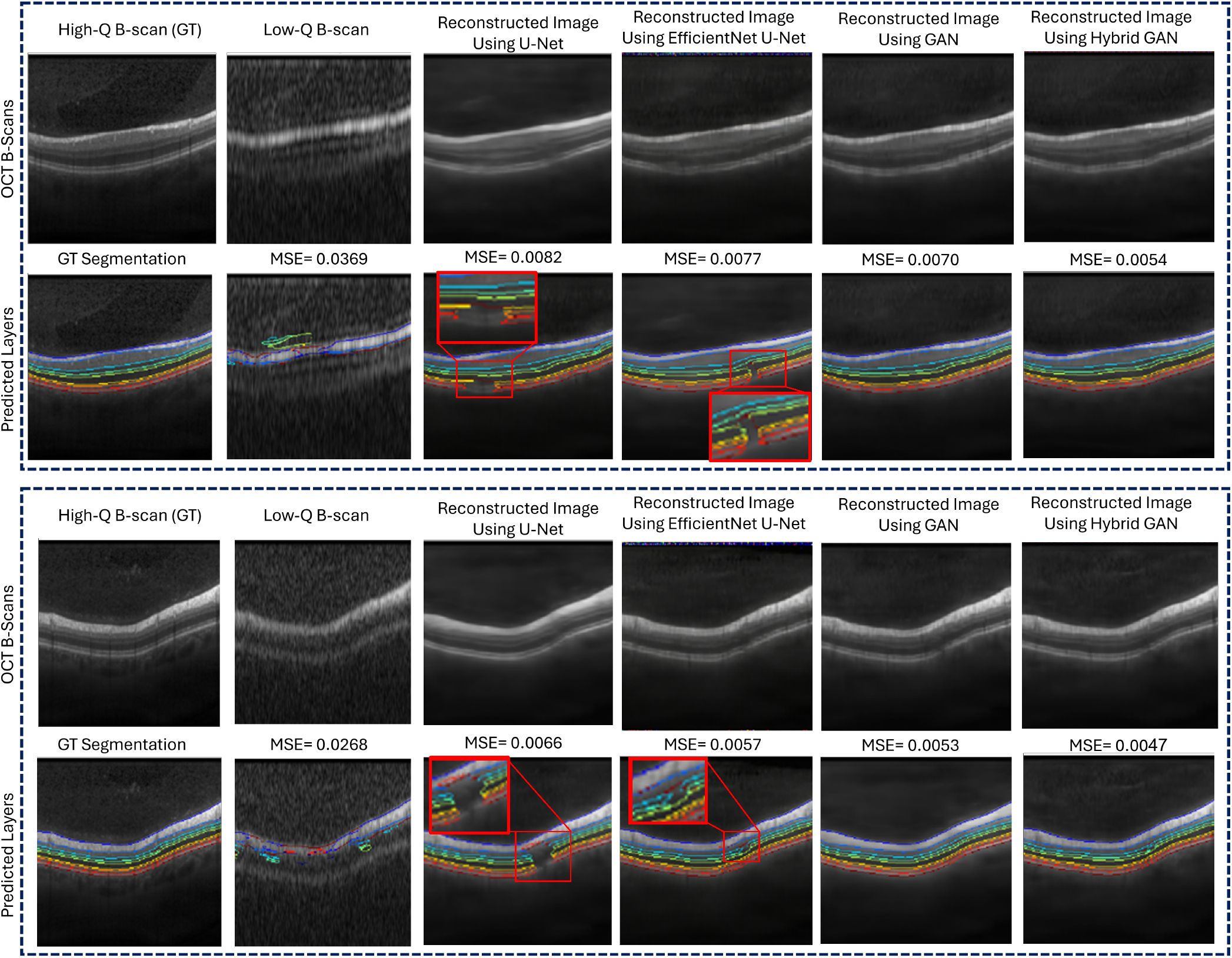
Qualitative comparison of retinal layer segmentation results obtained through the progressive model development strategy. For each sample, the first row shows, from left to right, the high-quality ground-truth OCT B-scan, the corresponding low-quality B-scan as the input for the models, and the enhancement results produced by the baseline U-Net, the EfficientNet-U-Net selected as the best-performing U-Net and autoencoder-based architecture, the GAN model using the winning EfficientNet-U-Net as the generator, and the proposed Hybrid Attention GAN (HAGAN). The second row presents the corresponding retinal layer segmentation results for each image. The baseline U-Net exhibits local discontinuities and fragmented layer boundaries, which are progressively reduced as the model architecture is refined. The final HAGAN yields the most consistent and anatomically coherent retinal layer segmentation.

Figure 7 further presents the learning curves of the four representative models considered above: the baseline U-Net, the EfficientNet-U-Net, the GAN model using the EfficientNet-U-Net as the generator, and the proposed HAGAN. The figure shows the training and validation loss trajectories for the U-Net–based models, along with the generator and discriminator losses for the GAN and HAGAN frameworks. As observed, the progressive refinement of the model architecture leads to improved convergence behaviour and lower loss values. In particular, the EfficientNet-U-Net converges faster and exhibits improved generalisation compared to the baseline U-Net. Building on this improvement, the GAN-based formulation further enhances the optimisation behaviour, achieving lower loss values and more stable convergence than the EfficientNet-U-Net alone. The proposed HAGAN yields additional improvements, demonstrating smoother convergence and reduced loss fluctuations for both the generator and the discriminator relative to the standard GAN, thereby underscoring the effectiveness of the proposed progressive design strategy.

**Figure 7:**
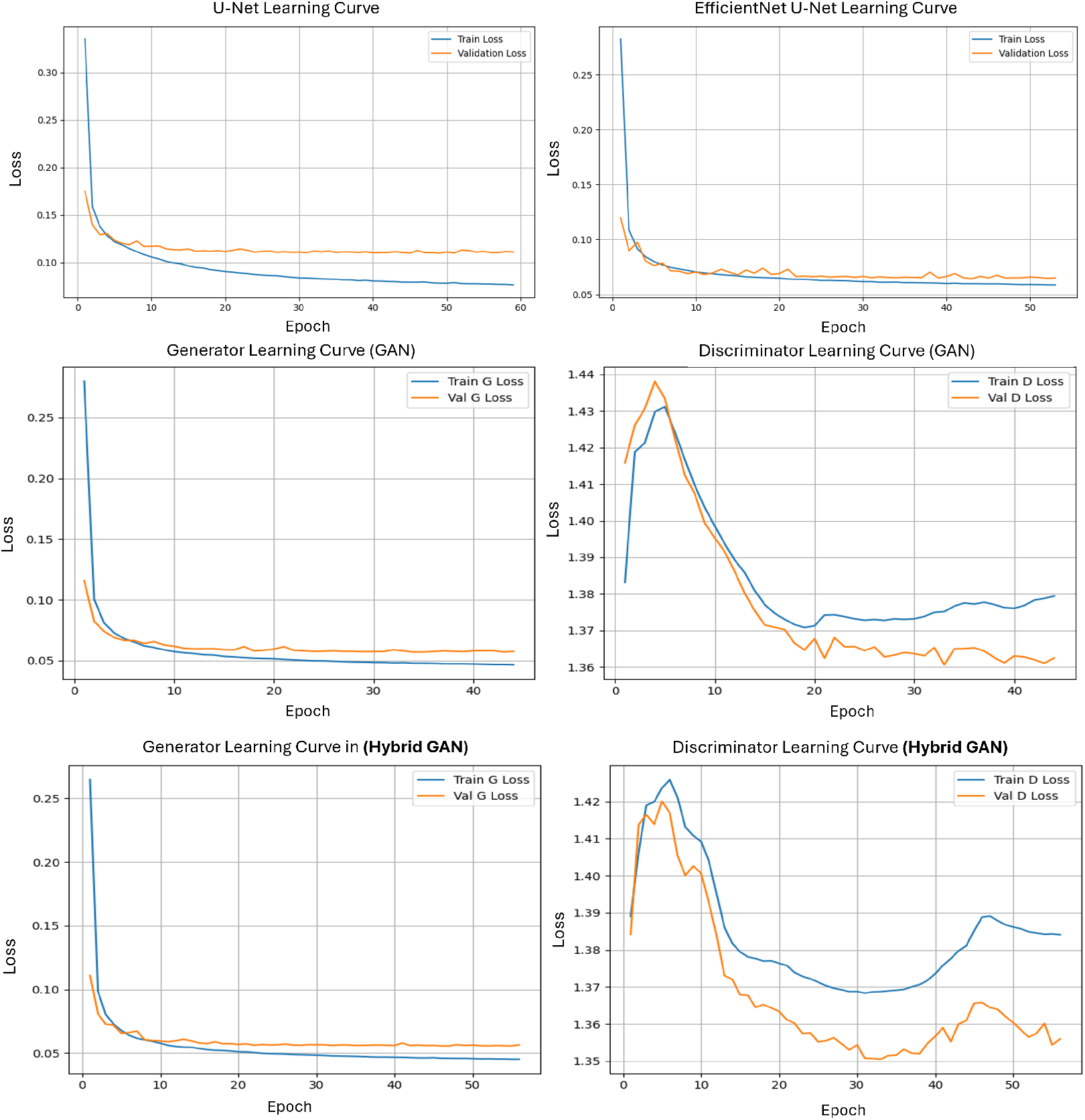
Learning curves of the four representative models considered in this study: the baseline U-Net, the EfficientNet-U-Net, the GAN model using the EfficientNet-U-Net as the generator, and the proposed Hybrid Attention GAN (HAGAN). The figure shows the training and validation loss trajectories for the U-Net–based models, as well as the generator and discriminator losses for the GAN and HAGAN frameworks. The EfficientNet-U-Net converges faster and generalises better than the baseline U-Net, while the GAN-based formulation achieves lower loss values and more stable convergence than the EfficientNet-U-Net alone. The proposed HAGAN further improves training stability, exhibiting smoother convergence and reduced loss fluctuations for both the generator and the discriminator compared to the standard GAN.

### 5.6. Comparison with State-of-the-Art OCT Denoising Models

To evaluate the proposed Hybrid Attention Generative Adversarial Network (HAGAN), we compared it with three representative GAN-based OCT enhancement methods as mentioned in the related work section: *SiameseGAN* [39], *SDSR-OCT* [40], and a *pix2pix* -based OCT denoising model with texture loss [41, 42].

As pretrained weights for *SiameseGAN* were publicly available, the released model was first evaluated directly on our test dataset to establish a baseline performance. To adapt the model to our data distribution, we performed domain adaptation through partial fine-tuning of the generator decoder. Specifically, we investigated several fine-tuning configurations by unfreezing different numbers of the final convolutional blocks of the decoder while keeping the remaining layers frozen. Three configurations were evaluated: fine-tuning only the final convolutional block, fine-tuning the last two decoder blocks, and fine-tuning the last three decoder blocks. This study allows us to assess how increasing the number of trainable decoder layers affects the adaptation performance while preserving the pretrained feature representations. In addition, the SiameseGAN architecture was also trained from scratch on our dataset to provide a fully comparable baseline. The evaluated fine-tuning configurations are illustrated in Figure 8. The figure provides a normalized comparison of SiameseGAN under different decoder fine-tuning strategies, including freezing all layers, progressively unfreezing the last one, two, and three decoder blocks, and training the model from scratch. For visualization, all metrics were normalized to [0, 1], and error-based metrics (MAE, MSE, LPIPS, and segmentation MSE) were inverted so that higher values consistently indicate better performance. The results demonstrate that increasing the number of trainable decoder layers improves SiameseGAN performance across both enhancement and segmentation metrics. Training from scratch yields a slight additional improvement compared with partial fine-tuning, while the proposed HAGAN model consistently achieves the best performance across all metrics.

**Figure 8:**
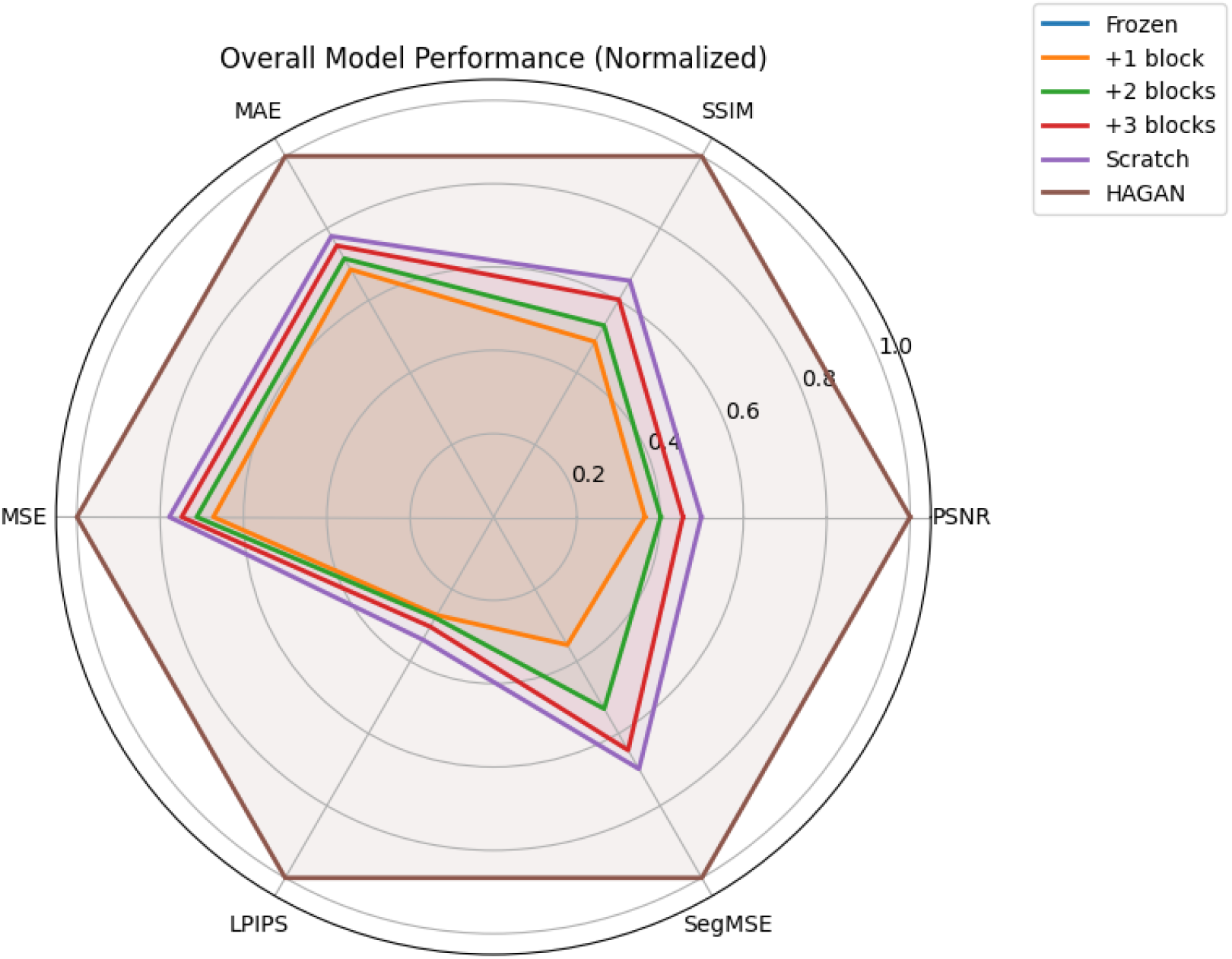
Normalized radar plot comparing the performance of *SiameseGAN* [39] under different fine-tuning configurations and the proposed Hybrid Attention GAN (HAGAN). The evaluated metrics include image enhancement quality (PSNR, SSIM, MAE, MSE, and LPIPS) as well as downstream retinal layer segmentation performance, measured by the average layer-wise segmentation mean squared error (SegMSE). All metrics were normalized to the range [0, 1] for visualization. Error-based metrics (MAE, MSE, LPIPS, and SegMSE) were inverted so that higher values consistently indicate better performance.

For *SDSR-OCT* and the *pix2pix* -based texture-loss GAN, pretrained weights and saved models were not publicly available. Therefore, both architectures were reproduced according to their original publications and trained from scratch using our training dataset.

A quantitative comparison between the proposed *HAGAN* model and all baseline methods is reported in Table 7. In this comparison, all models were trained from scratch using the same training dataset and evaluated on the same test set, ensuring identical experimental conditions and a fair comparison. The results show that *HAGAN* consistently achieves the best performance across all enhancement and segmentation metrics, indicating superior reconstruction quality and improved preservation of retinal structural details compared to existing GAN-based approaches.

**Table 7:** Quantitative comparison of state-of-the-art GAN-based OCT image enhancement models trained from scratch using the same training dataset and evaluated on the same test dataset. Lower values indicate better performance for metrics marked with *↓*, while higher values indicate better performance for metrics marked with *↑*.

| **Metric** | **pix2pix [41, 42]** | **SDSR-OCT [40]** | **SiameseGAN [39]** | **HAGAN (Proposed)** |
| --- | --- | --- | --- | --- |
| **Enhancement Metrics** |  |  |  |  |
| MAE↓ | 0.02870 | 0.0261 | 0.0385 | **0.0181** |
| MSE↓ | 0.00239 | 0.00190 | 0.00445 | **0.00090** |
| PSNR↑ | 26.78 | 28.01 | 24.35 | **30.83** |
| SSIM↑ | 0.7662 | 0.7815 | 0.7280 | **0.8537** |
| LPIPS↓ | 0.1723 | 0.2502 | 0.412 | **0.1542** |
| **Segmentation Performance (Average Boundary MSE ↓)** |  |  |  |  |
| Avg | 0.0086 | 0.0049 | 0.00991 | **0.0029** |

## 6. Discussion

The experimental results demonstrate the effectiveness of the proposed HAGAN framework in addressing the unique challenges of remote and patient-operated OCT image enhancement. Through a systematic comparison of multiple encoder backbones within a U-Net architecture [29, 34, 35], we identified EfficientNet-B1 as the most suitable encoder for preserving fine retinal details while maintaining high-level contextual awareness. This finding confirms that encoder backbone selection plays a critical role in balancing reconstruction fidelity and representational capacity in medical image enhancement tasks.

Ablation studies further highlight the importance of both skip connections and transfer learning. Models with frozen low-level encoder layers leveraged generic features learned from natural images while fine-tuning deeper layers for OCT-specific structures, yielding improved generalization and reduced training cost. This strategy provided a strong foundation for subsequent GAN-based enhancement.

The introduction of adversarial learning addressed key limitations of reconstruction only models, which tend to produce over-smoothed outputs that obscure subtle anatomical boundaries. The VGG-based discriminator encouraged the recovery of sharper textures and improved perceptual realism, as reflected by consistent gains in LPIPS [48]. However, comparisons against a no-attention GAN baseline representative of conventional adversarial OCT enhancement methods [37] indicate that adversarial training alone is insufficient to simultaneously optimize local detail refinement and global structural coherence.

Attention mechanisms proved essential for resolving this trade-off. Comparisons with AG-only (AGGAN) and SA-only (SAGAN-style) [43] generators demonstrate that attention gates primarily enhance local structural fidelity by suppressing irrelevant encoder features, while self-attention improves global consistency by modeling long-range spatial dependencies. Notably, neither mechanism alone achieved optimal performance across all evaluation criteria. The hybrid AG+SA configuration consistently outperformed both isolated variants, delivering the best balance between pixel-level fidelity, perceptual quality, and segmentation-based accuracy. These results highlight the complementary roles of local and global attention in structure-preserving OCT image enhancement.

Additionally, comparison with recent GAN-based OCT restoration approaches, including SiameseGAN [39], SDSR-OCT [40], and pix2pix-based methods [41, 42], indicates the superior performance of the proposed HAGAN framework. This improvement can be attributed to the fact that, while these models primarily address noise reduction or resolution enhancement, remote OCT imaging may introduce additional artifacts and acquisition irregularities. The proposed HAGAN framework addresses this limitation by integrating adversarial learning with hybrid attention mechanisms, enabling simultaneous suppression of noise and mitigation of imaging artifacts while preserving clinically relevant retinal structures.

Importantly, model performance was evaluated not only using standard image quality metrics but also through a clinically relevant downstream task: retinal layer segmentation. Consistent improvements in segmentation accuracy across nine retinal layers indicate that HAGAN enhances anatomically meaningful information rather than merely improving visual appearance. This task-based validation provides stronger evidence of clinical relevance compared with evaluations based solely on perceptual or fidelity metrics. Robustness to severe image degradation was further investigated under progressively increasing noise and artifact levels. As shown in Subsection 5.4.2, although the reconstruction task becomes increasingly challenging as the degradation severity grows, HAGAN remains capable of recovering meaning-ful retinal structures and preserving key anatomical boundaries. The gradual decline in quantitative metrics further indicates that the model maintains stable behavior even under heavily corrupted inputs, highlighting its robustness to severe noise and artifact conditions.

While the results are promising, several limitations should be acknowledged. First, the training data were generated using a simulation framework intended to approximate the characteristics of a low-performing OCT device. Although simulation enables controlled and reproducible experimentation, the simulated images originate from high-quality hospital OCT scans acquired using a different imaging system (e.g., Heidelberg devices), with noise and artifacts synthetically introduced. Despite visual similarities in noise and artifact patterns, inherent differences persist between simulated and real remote patient-operated acquisitions, including spatial resolution, sampling density (e.g., number of A-scans), and other device-specific imaging characteristics. This domain discrepancy underscores the need for validation using real-world remote OCT data. As demonstrated in Subsection 5.6, pretrained models can be adapted to previously unseen data through fine-tuning, providing a straightforward mechanism for adjusting the model to a new data distribution. While fine-tuning represents the simplest and most practical approach for such adaptation, more advanced domain adaptation techniques could also be employed to further reduce the domain gap between simulated training data and real-world OCT images, thereby improving the generalization of models trained on simulated data to remote acquisitions.

Finally, further investigation is required to assess generalization across diverse patient populations and a broader range of pathological conditions, ensuring robust performance in real-world applications.

## 7. Conclusion

In this paper, we introduced HAGAN, a Hybrid Attention GAN designed to enhance low-quality remote OCT images. Building upon an EfficientNet-U-Net backbone, HAGAN integrates both attention gates and self-attention within a GAN framework, guided by a comprehensive multi-objective loss function. Extensive experiments demonstrate that the proposed model consistently outperforms both baseline architectures and recent sate-of-the-art GAN-based OCT restoration methods, including SiameseGAN, SDSR-OCT, and pix2pix-based approaches, producing reconstructions with improved pixel-level accuracy, perceptual quality, and anatomical fidelity while effectively addressing the complex degradations characteristic of a significantly reduced-performance OCT imaging systems.

Beyond conventional metrics, we validated the clinical relevance of HAGAN by assessing its impact on retinal layer segmentation. The observed improvements in segmentation accuracy confirm that the enhanced images retain diagnostically meaningful structures, bridging the gap between algorithmic performance and clinical applicability.

In summary, HAGAN combines local and global attention with adversarial learning to enable robust enhancement of remote OCT images, supporting reliable and frequent retinal monitoring outside specialized clinical settings. By improving image quality from patient-acquired scans, the approach has the potential to reduce dependence on in-clinic imaging and facilitate timely assessment of disease progression, particularly benefiting older and mobility-limited patients. While this study is based on physically-accurate simulated data, future work will focus on validation with real-world device data, integration of domain adaptation strategies to bridge the simulation–reality gap, optimization for real-time deployment, and extension to pathological datasets to support clinical translation.

## Data Availability

All data produced in the present study are available upon reasonable request to the authors

## Code Availability

The implementation of the proposed HAGAN framework is publicly available to support reproducibility and further research at: https://github.com/royaarian101/HAGAN.

## Acknowledgment

This work was supported by the Engineering and Physical Sciences Research Council [grant number EP/X525546/1] through the Impact Acceleration Account (IAA), for the project entitled “Enhancing Healthcare with AI-Based Home Monitoring for Retinal Diseases” (Project ID 1709598:).

